# Age separation dramatically reduces COVID-19 mortality rate in a computational model of a large population

**DOI:** 10.1101/2020.05.27.20111955

**Authors:** Liron Mizrahi, Shani Stern

## Abstract

COVID-19 pandemic has caused a global lock down in many countries throughout the world. Faced with a new reality, and until a vaccine or efficient treatment is found, humanity must figure out ways to keep economy going on one hand, yet keep the population safe on the other hand, especially those that are susceptible to this virus. Here we use a network simulation, with parameters that were drawn from what is known about the virus, to explore 5 different scenarios of partial lock down release. We find that separating age groups by reducing interactions between age groups, protects the general population and reduces mortality rates. Furthermore, addition of new connections within the same age group to compensate for the lost connections outside the age group, still has a strong beneficial influence and reduces the total death toll by 66%. While complete isolation from society may be the most protective scenario for the elderly population, it would have an emotional and possibly cognitive impact that might outweigh its benefit. We therefore propose creating age-related social recommendations or even restrictions, thereby allowing social connections but still strong protection for the older population.

## Introduction

COVID-19 pandemic started late December of 2019 with a mysterious pneumonia in Wuhan, China and was declared a global pandemic by the World Health Organization (WHO) on March 11, 2020. The disease has quickly spread throughout 6 continents and over 210 countries. COVID-19 causes a respiratory disease and is considered to be much more contagious than the influenza ^1^. Common symptoms include fever, cough, fatigue, shortness of breath, and loss of smell ^2-4^. COVID-19 related complications include pneumonia, and acute respiratory distress syndrome that may develop into a severe respiratory failure, septic shock and death ^5,6^. In addition to being more contagious than influenza, COVID-19 has longer incubation periods compared to influenza. During the incubation period the patients may be contagious ^7-10^. Reports present incubation periods with a mean of 5-6 days ^7,11^, during which the patients are contagious. Additionally, the mortality rate from COVID-19 disease is higher than mortality rate from influenza complications ^12^. These conditions instigated a rapid spread of the disease, causing over 100 countries to declare lock downs and curfews and causing an estimated global economy loss of 1 trillion dollars in 2020^13^. Till this date almost 6 million people were tested positive to COVID-19 and more than a 350,000 people died from the virus-related complications.

Mortality rate from COVID-19 is strongly age biased, affecting the older population to a much greater extent ^14-16^. In fact, it is thought that younger population is usually asymptomatic or experience mild symptoms, even when infected with the virus ^10,17^. For those symptomatic patients the incubation period is the same regardless of the age. Recovery is reported to be 28±14 days ^18^.

When the global lock down is released, a slow release of the population back to their daily routine will occur. There is thought to be a psychological toll ^19-21^ for the isolation of the population. Since COVID-19 is lethal mostly to the elderly population, suggestions of reopening the curfews, but keeping the elderly population isolated have been suggested ^22-24^. The psychological, emotional and even cognitive impact may be stronger on this population, as social interactions is known to be essential for preventing cognitive and physical decline ^19,23,24^. If therefore there could be a solution where social interactions would not be prevented for the elderly population but keeping the population at a low risk from the virus, this may be a preferred solution to this population.

The lower chances of the older population to be asymptomatic or to present mild symptoms compared to younger population, means that a COVID-19 older patients are contagious for shorter periods of time compared to the younger patients. Asymptomatic (or weakly symptomatic) patients may be contagious all the way until full recovery (28±14), whereas a symptomatic patient will be contagious for 6.4±2.3 days. This makes a young individual as a stronger candidate of infecting others compared to an older individual. Here we used these assumptions to derive a model of a large population to see what happens if we allow social interactions in the elderly population, but only among their own age group. This type of restriction will allow for these social interactions that are so important for the elderly age group. The model confirms that there would be a drastic reduction in mortality rate compared to allowing social interactions with other age groups (between 66-88%, depending on the scenario), even when we keep the total number of connections to be the same by adding new connection within the elderly population for every lost connection with the younger population.

## Methods

### Population and network connectivity

We simulated a population of 50,000 individuals using network theory and a Erdős-Rényimodel network with degree distribution of 15 ^25^. We separated the population into 4 age groups: 0-14, 15-34, 35-54, 55+. The distribution of the population among the different age groups was chosen to be similar to the population distribution in Israel and is presented in Figure 1.

**Figure 1.**
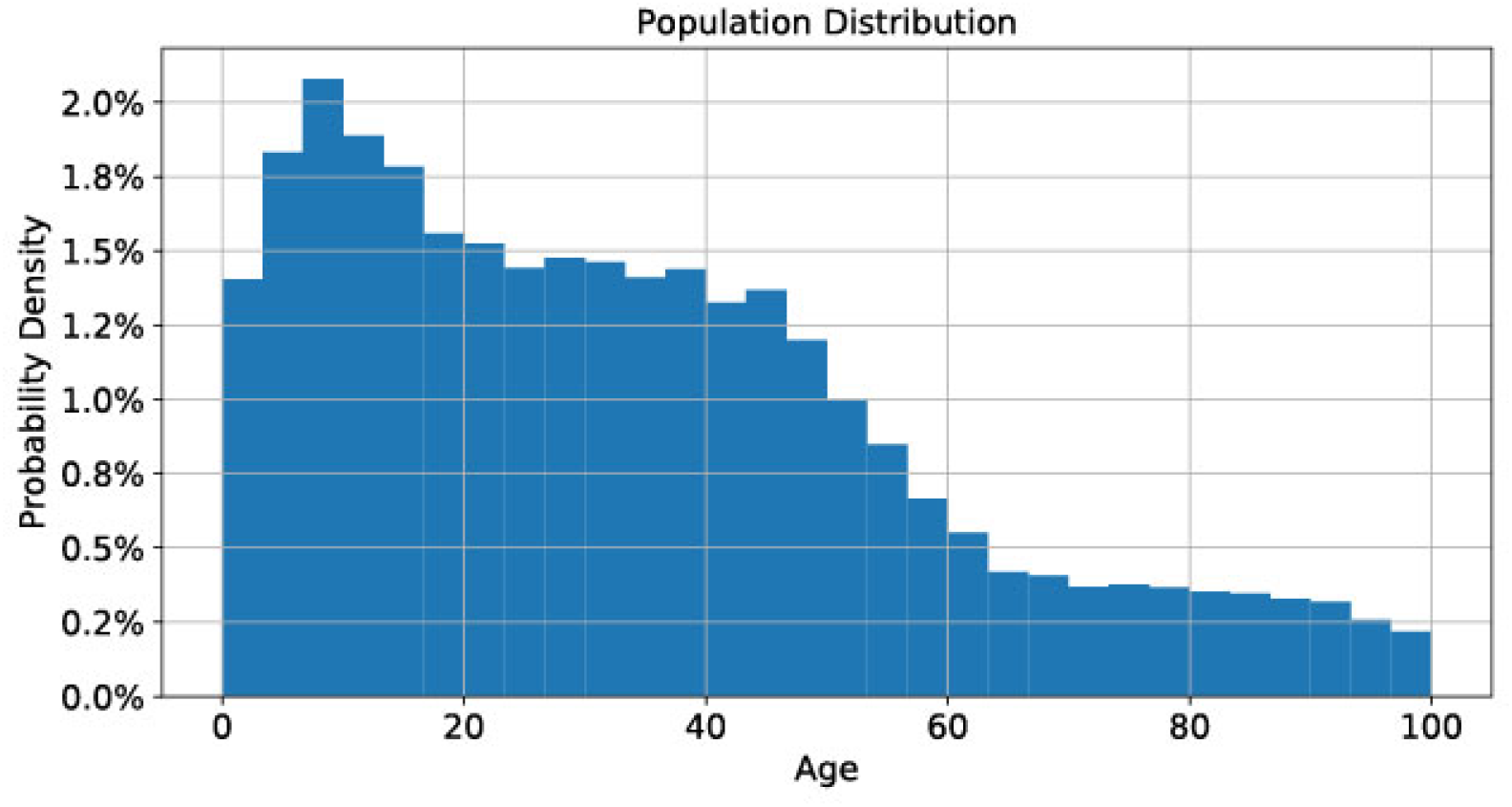
The distribution of the population by their age.

We chose the size of the families (or household) to be normally distributed to be 4±1. Families were constructed in the following manner. After randomly grouping the entire population into families using a Normal distribution with a mean of 4 and a standard deviation of 1, the family members were filled in according to the size of the family; If the family size was larger than 2, a pair of parents aged 20-60 were randomly taken from the population pool and the remaining members of the family were randomly selected with an age of 0-20. If the family had only 2 members, 2 adults with an age of 55-100 were randomly selected, and if the family size was 1, one adult with an age of 20-100 was randomly selected from the general population. These constraints resulted in the following family sizes according to the age groups: 4.5±0.94 for the first age group, 4.3±0.93 for the second group, 4.1±0.90 for the third age group, 1.9±1.24 for the fourth. A plot of the population distribution over the age is plotted for the different age groups in Figure 2.

**Figure 2.**
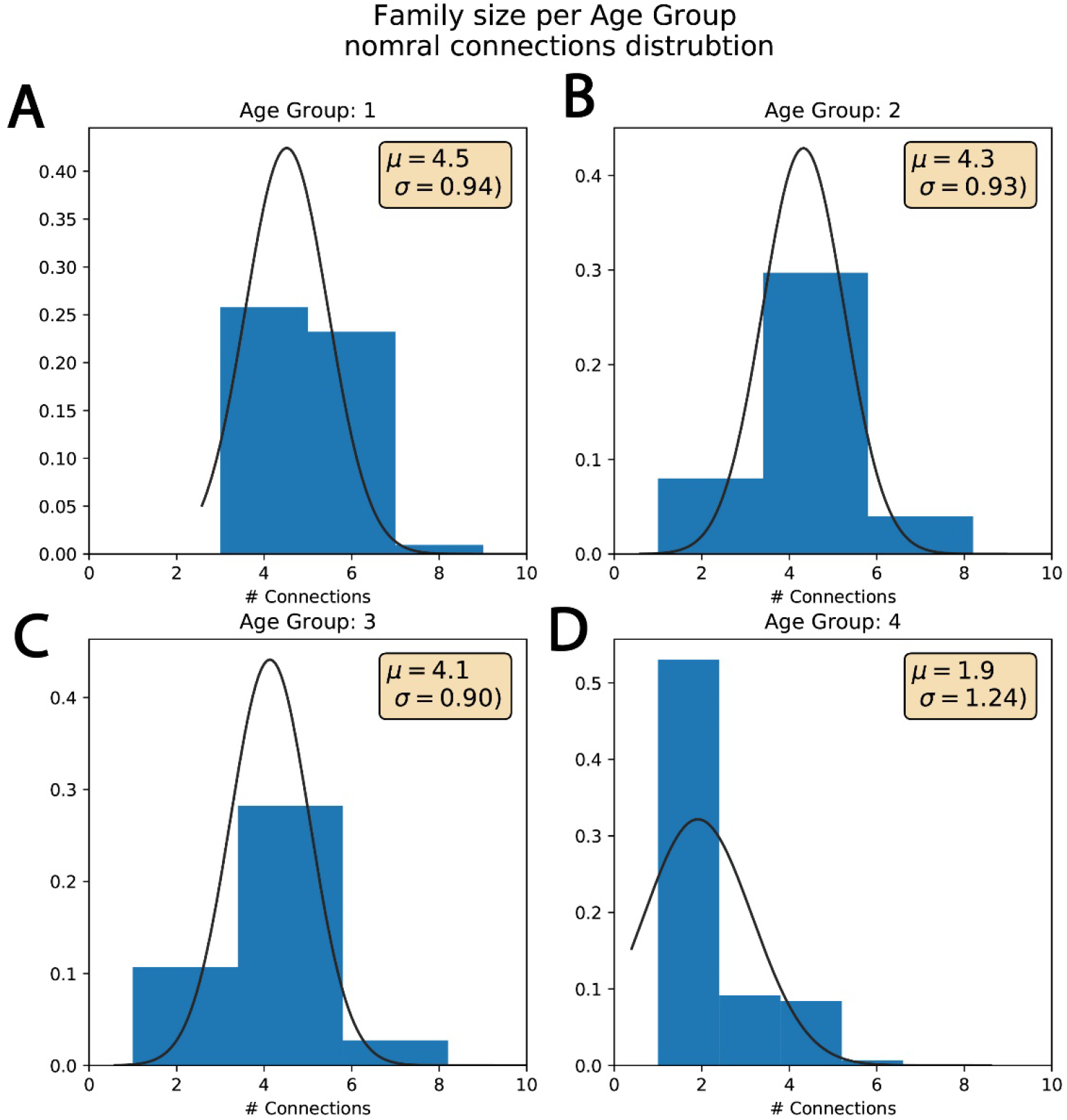
The distribution of family size by age group. A. The family size distribution in age group 1 (0-14). B. The family size distribution in age group 2 (15-34). C. The family size distribution in age group 3 (35-54). D. The family size distribution in age group 3 (55+).

Due to the family members selection, the connectivity level in the different age groups also varied and consisted of 15±3.82 connections for the first age group, 15±3.79 connections for the second age group, 15±3.82 for the third age group and 14±3.80 connections for the fourth age group. Figure 3A shows the distribution of the number of connections according to the different age groups (these conditions will later be defined as state 1). Figure 4 presents example connections within the general 50000 individuals’ population of 3 individuals (Fig. 4A) and of 30 individuals (Fig. 4B).

**Figure 3.**
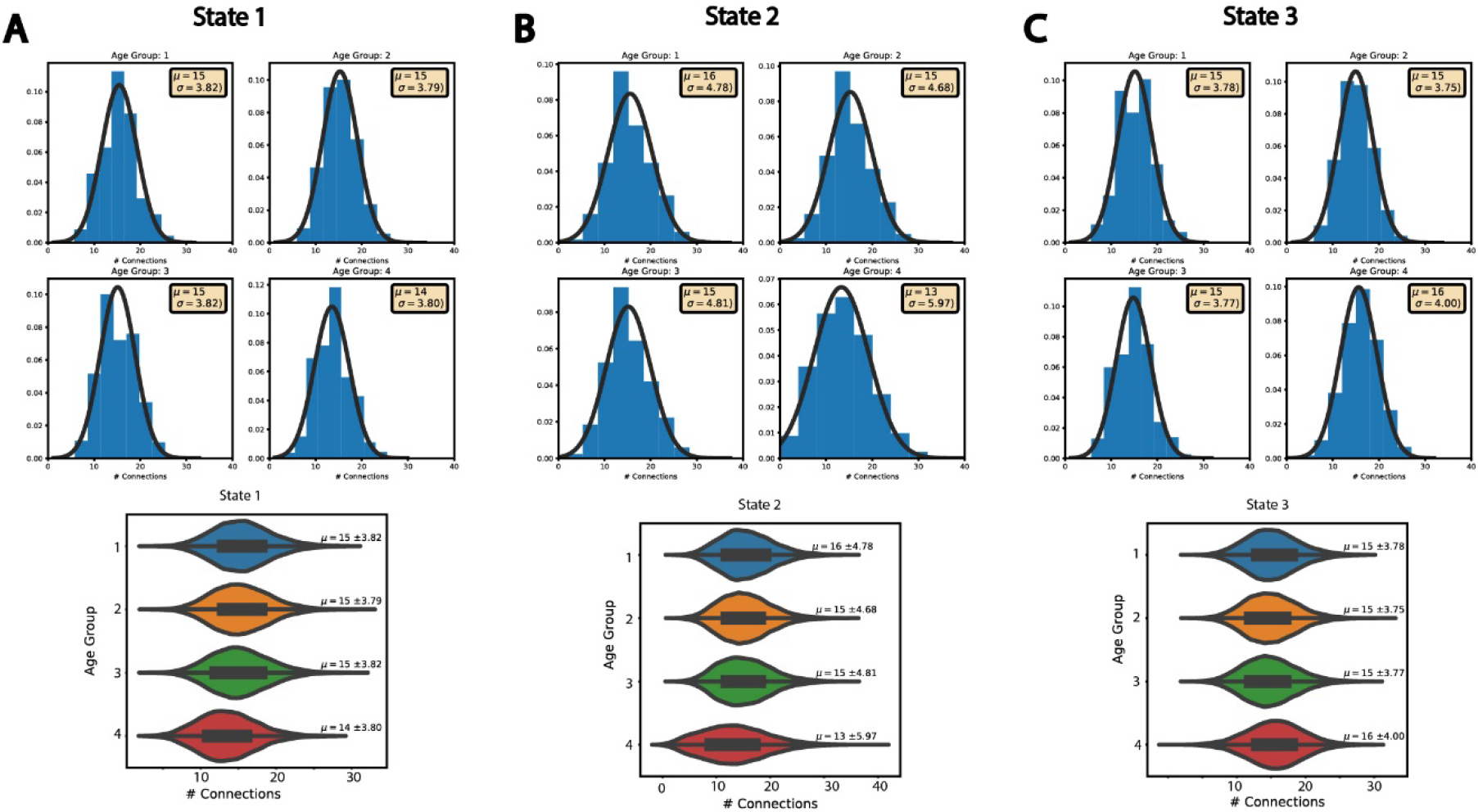
The distribution of the number of connections per individual in the different age groups in states 1-3 shows similar connectivity. A. The distribution of the connections in state 1 in the different age groups. B. The distribution of the connections in state 2 in the different age groups. C. The distribution of the connections in state 3 in the different age groups. The number of connections did not change much between the states, indicating that disease evolution in these states changes mainly due to the age separation and not due to changes in the connectivity of the network.

**Figure 4.**
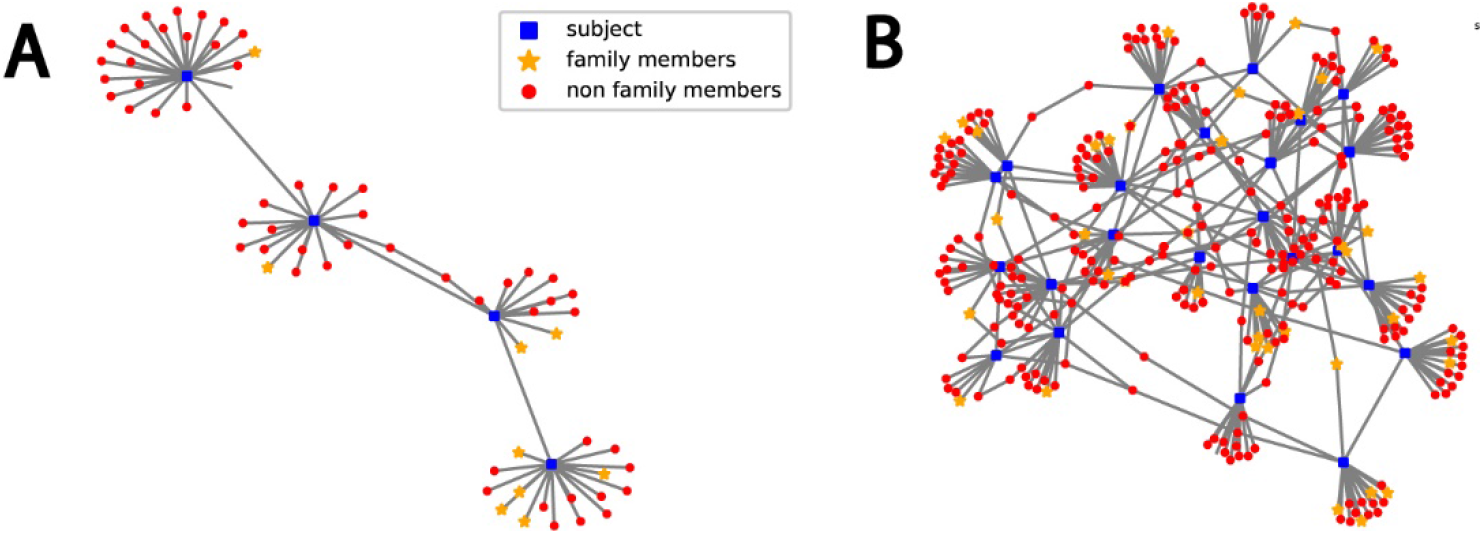
Example connections within the population. A. Example of connections between 3 subjects. B. Example connections between 30 subjects.

### Infection

The model assumes different infection rate at different scenarios, but what is common is that once an individual becomes symptomatic, he is assumed to be under full quarantine and is therefore removed from the network. The highest infection rate occurs within the family and is 10% daily; each day each infected individual will infect another individual within his family with a probability of 10%. The infection rate in public is reduced to 1%, if people keep social distancing, thus reducing dramatically the infection rate.

Once infected, symptoms may appear in all age groups under a different probability. There is a higher probability to be asymptomatic (or presenting mild symptoms) for younger people. We used 80% chance to be asymptomatic for the 0-14 age group. 60% chance to be asymptomatic for the 15-34 age group, 40% chance for the 35-54 age group, and 20% chance for the 55+ age group. These numbers were based on several reports ^17,26,27^. For those patients that will become symptomatic, the number of days until becoming symptomatic has a Weibull distribution with a mean of 6.4 days and standard deviation of 2.3 days to start being symptomatic ^11^, and this does not vary between the age groups.

Mortality rates varies dramatically among age groups and is 0% for 0-14, 0.15% for 15-34, 1% for 35-54 and 24% for ages 65+, similar to what was reported in^14^. Once showing symptoms the recovery rate (for those who recover) is 28±14 days (Normal distribution) in all age groups^18^.

## Results

We defined 5 states. The first state complies with all the above assumptions and with no other restrictions. Supplemental Video 1 shows the spread over time of COVID-19 demonstrating graphically the connections and infections in individuals in a population of 10,000 people for state 1. Blue dots represent susceptible individuals, orange dots represent exposed individuals, red dots represent infected individuals, and black dots are individuals who dies from COVID-19. The shape of the dots marks the age groups (a circle 0-14, a triangle 15-34, a square 35-54 and an x for 55+). We next wanted to test how age separation changes infection and mortality rates in the population. For this, we defined 4 more states with different scenarios of age separation. In state 2 we did not allow interactions between the different age groups, such that the interactions among family members remained without a change, but all other connections were eliminated. This reduced the connectivity of the network. Since we were interested to see the effect of the age separation and not the effect of reducing the connectivity, we added in state 2 new random connections within the same age group to replace any connection that was eliminated. The new number of connections for each group resembles the original conditions and is shown in Fig. 3B and is similar to the original number of connections (the family size distribution remains since we had not changed the family connections).

We next defined a more plausible constellation of the network; In state 3 we grouped the 3 age groups 0-14, 15-34, and 35-54, and did not allow connections with the elderly group of 55+ (except if they are in the same household). Such restrictions also reduced the number of overall connections, and we therefore added random connections within the same age group to any connection that was eliminated between the age groups. Figure 3C presents the new distribution of connections per individual in the different age groups, and the number of connections is similar (or greater) than the original number of connections.

Since it is also reasonable to assume that restrictions of connections between age groups may reduce network connectivity, we added state 4 in which we keep all the connections, but reduce the connectivity between age groups, by reducing by half the infection rates to 0-5% between age groups. We similarly defined state 5, where we completely diminish connections between different age groups (but without adding new connections).

Figure 5 presents the simulation results over a 250 days’ time period in a population of 50000 individuals by the categories of susceptible, exposed (was in contact with a COVID-19 positive person), infected, recovered and deceased for the entire population pooled together in the 5 different states. We expanded the plots to see in more detail the infection and death rates. Figure 6A presents the infection over the 250 days for the 5 different states. Figure 6B similarly shows mortality from the virus over the course of 250 days in the different states. Further comparison of infection and mortality rates (Figure 6A, 6B respectively) shows dramatic infection rates changes, and most importantly lower mortality in the different states (2-5) compared to state 1. This shows that reducing the interaction between age groups decreases mortality, even when keeping the number of interactions between individuals constant or even increasing this number. Separating just the older group (55+) from interacting with the other age groups (but keeping interactions within the same household interactions, state 3) reduces the overall mortality rate in the population by 66% (!). This decrease becomes even more pronounced when not adding new connections within the same age group (state 5) with a reduction of 88% in mortality rate in the overall population. Using this strategy in a real-world scenario, would probably result in a reduction that is somewhere between these 2 states, since some interactions will be replaced (such as seats in the theater), but some will be eliminated (such as meeting distant relatives).

**Figure 5.**
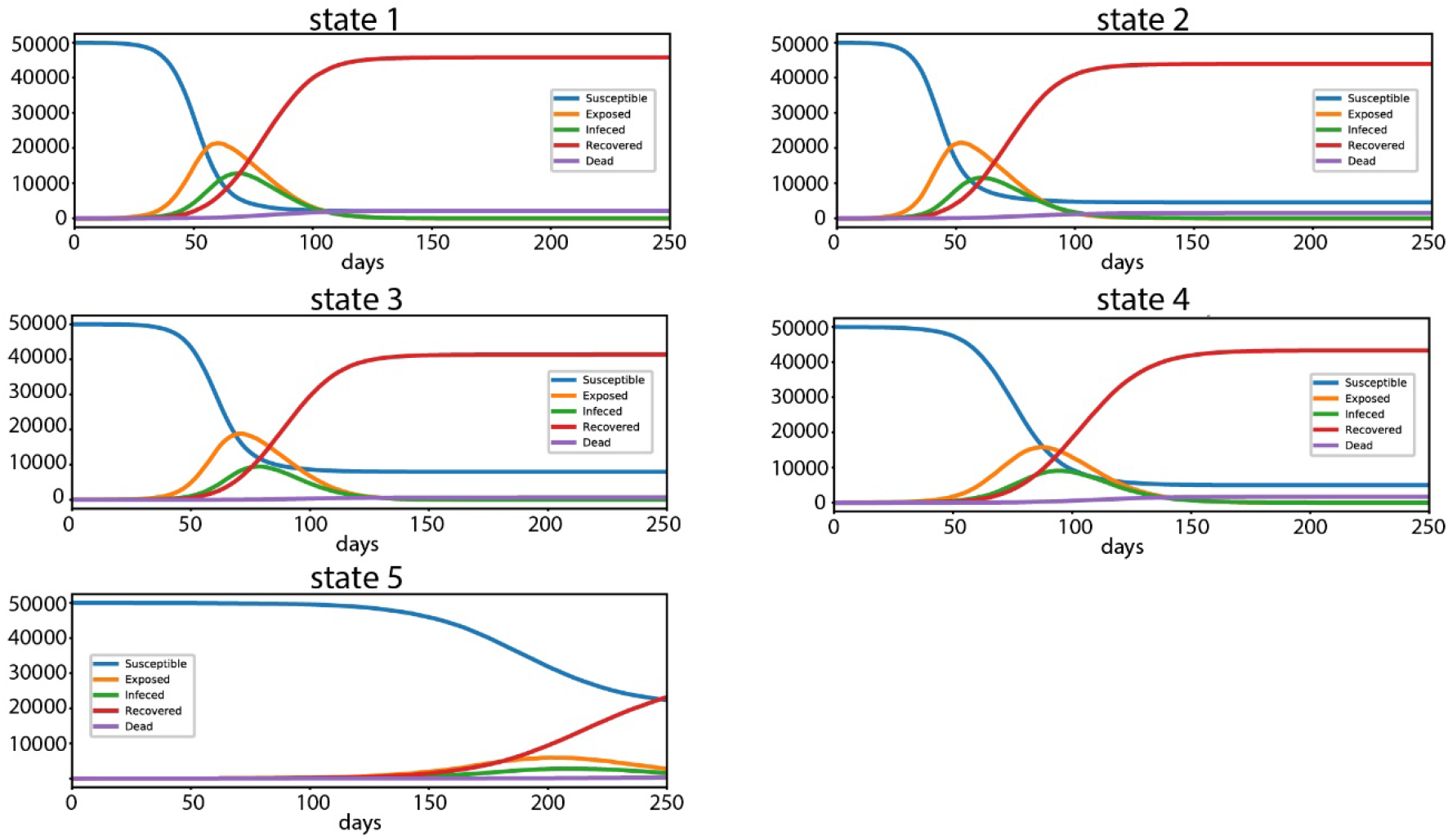
The total number of susceptible, exposed, infected, recovered and deceased individuals in the entire population of 50000 people over a period of 250 days in the different states. A-E. for state1-state5 respectively.

**Figure 6.**
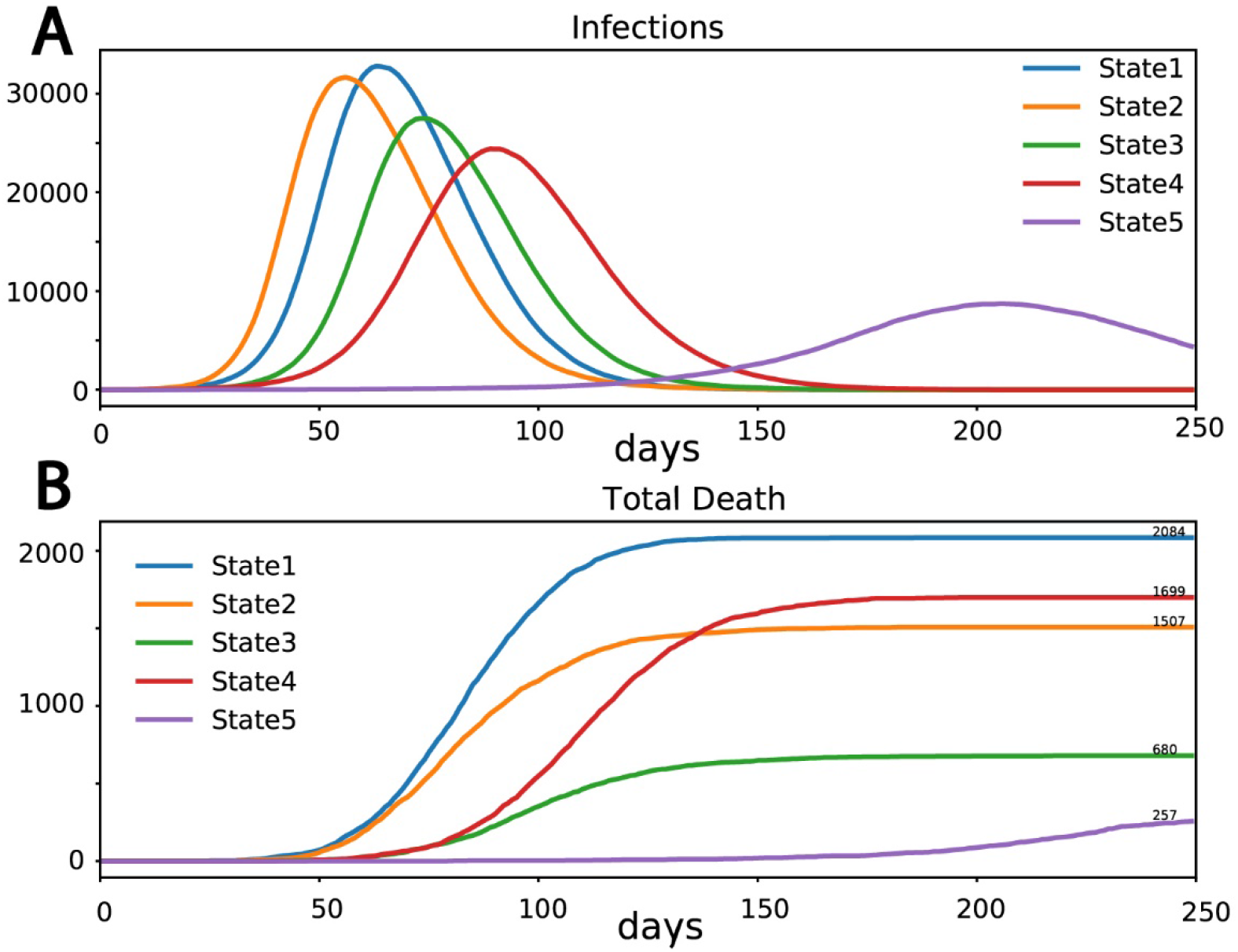
A. Total number of infections in the entire population for the different states. B. Total number of deceased in the entire population in the different states. While in state 1 2084 individuals died, in state 2 only 1507 individuals died, in state 3 only 680 individuals died. In state 4 1699 individuals died and in state 5 only 257 individuals died.

Supplemental Video 2 presents the spread of the disease in states 1-4. The color and shape schemes are the same as in Supplemental Video 1. Supplemental Video 3 similarly shows the same disease evolution as Supplemental Video 2, but the blue dots were removed for clearer graphs. The clear reduction of death rate is exhibited in the new states.

## Discussion

COVID-19 caught the world unprepared and has infected (to this day) over 4 million people and has cost over a quarter of a million lives. The pandemic has caused a global lock-down that in turn caused a huge economic burden. Releasing the lock down needs to be under controlled conditions, being extremely careful not to cause a “second wave” ^28,29^. The most susceptible population is the elderly population, and there have been suggestions of releasing the general population, keeping the elderly quarantined. However, social interactions are known to be extremely important in the elderly population ^30,31^, and such conditions may backfire, causing mental and emotional deterioration in this population.

In this study we tested several states that limit the interactions between people in different age groups, and we showed that all these scenarios reduce mortality rate. In the first two states, we kept the interactions of family members within the same household, and eliminated connections outside of the age group, yet keeping the network connectivity (by adding more connections within the same age group). This drastically reduces the overall death toll by 66%. The reasoning for the improvement in the overall mortality is that younger people have a higher probability of being asymptomatic (or weekly symptomatic). Asymptomatic people usually keep on their social interactions, infecting many people for a very long time. Older people have a much lower probability of being asymptomatic. This means that they are usually infectious for only ~5-6 days (the asymptomatic period is assumed to have a Weibull distribution with an average of 6·4 days). At this point they usually know that they are sick and keep quarantined. Therefore, an older individual is less likely to spread the disease.

In the other two states, we reduced the interactions (state 4) or completely diminished the interactions (state 5) between the different age groups. This may be a reasonable assumption, since not all connections between age groups will be replaced by connections within the age group (such as connections with distant family). State 5 where all connections outside the age group is eliminated, reduces mortality rate by 88%. A real scenario will probably be some compromise between state 3 and state 5, since some connections will indeed be replaced (for example seating in a resultant, and some will be lost, such as meeting distant family members).

It is important to note that although age separation is difficult generally, it is relatively easy to find microenvironments in which age separation is possible and will enable the older population to maintain social connections while reducing infection and death rates in these populations. Such microenvironments may include grocery shops, theaters or airplane rides, and would allow important social interactions for the elderly population. Overall, our study shows that age separation is extremely beneficial and can be imposed in an important intermediate period until resuming normal life when finding of a cure or a vaccine to COVID-19.

## Data Availability

no data to report

## Supplementary

### Video1

A video demonstrating the spread of COVID-19 throughout a 10,000 individuals’ population for state 1. A. The entire population. Blue are susceptible individuals, Orange are exposed individuals, red are infected individuals and black are deceased. The first age group (0-14) is marked in circles, the second age group (15-34) is marked in triangles, the third age groups (35-54) is marked in squares, and the fourth age group (55+) is marked in x-s. The video shows the evolution of the disease in state 1.

### Video 2

A video demonstrating the spread of COVID-19 throughout a 10,000 individuals’ population for states 1-4. A. Blue are susceptible individuals, Orange are exposed individuals, red are infected individuals and black are deceased The first age group (0-14) is marked in circles, the second age group (15-34) is marked in triangles, the third age groups (35-54) is marked in squares, and the fourth age group (55+) is marked in x-s.

### Video 3

Presenting the same conditions as Supplemental video 2, but the blue dots have been removed for a clearer observation.

## Competing interests

Dr. Stern has nothing to disclose.

## Notes

### Competing Interest Statement

The authors have declared no competing interest.

### Funding Statement

No external funding was received

### Author Declarations

Committee on Research with Human Participants of University of Haifa

